# A Multidirectional Two-tube Method for Chemical Pleurodesis could improve distribution of the Sclerosing Agent within the Pleural Cavity

**DOI:** 10.1101/2021.04.06.21254972

**Authors:** Eoin Campion, Saad I. Mallah, Maimoona Azhar, Dara O’Keeffe, Aamir Hameed

**Affiliations:** School of Pharmacy and Biomolecular Sciences, RCSI University of Medicine and Health Sciences, Dublin 2, Dublin, Ireland; School of Medicine, RCSI University of Medicine and Health Sciences – Bahrain; Graduate Entry Medicine, School of Medicine, RCSI University of Medicine and Health Sciences, Dublin 2, Dublin, Ireland; Department of Surgery, St. Vincent’s University Hospital, Dublin 4, Dublin, Ireland; Department of Surgical Affairs, RCSI University of Medicine and Health Sciences, Dublin 2, Dublin, Ireland; Tissue Engineering Research Group (TERG), Department of Anatomy and Regenerative Medicine, RCSI University of Medicine and Health Sciences, Dublin 2, Dublin, Ireland; Trinity Centre for Biomedical Engineering (TCBE), Trinity College Dublin (TCD), Dublin, Ireland

**Keywords:** Malignant pleural effusion, chemical pleurodesis, talc slurry, pleural cavity, chest tube

## Abstract

**Introduction:** Malignant pleural effusion (MPE) affects approximately 200,000 people in the United States per annum. Chemical pleurodesis is a recommended first line treatment in the management of MPE, however, success rates as low as 43% has been reported. A bedside chemical pleurodesis can cost up to $11,224 and an estimated inpatient annual expenditure of more than $5 billion in the US alone. This study aims to assess the distribution of the talc slurry within the pleural space using cadaveric models and to determine the force required to push the talc slurry though a 14 Fr chest tube.

**Materials and Methods:** The force required to administer the talc slurry through a 14 Fr chest tube was tested using a Zwick/Roelle Z005 mechanical tester. Talc slurry distribution within the pleural cavity was assessed by direct visualisation following administration to the cadaveric models using single and multidirectional two-tube methods.

**Results:** Maximum force required to push the talc slurry though a 14 Fr chest tube was 11.36 N +/- 2.79 N. Distribution of the talc slurry within the pleural cavity was found to be poor with a single tube method. Multidirectional two-tube method of administration showed more even distribution.

**Conclusion:** The experimental multidirectional two-tube method results in wider distribution of the talc slurry within the pleural cavity and could further improve success rate of the talc pleurodesis.

## Introduction

Malignant pleural effusion (MPE) is a common complication in neoplastic disease, with growing prevalence. In the US, it is estimated that by the end of 2020, there would be 228,820 new lung cancer cases, 279,100 new breast cancer cases, 85,720 new lymphomas and 333,680 new gastrointestinal cancer cases [1]. MPE is a common complication in these patients. Yearly, there are over 150,000 new cases of MPE in the US alone [2]. Median survival time after diagnosis of MPE is 3 to 12 months depending on the stage and primary site of malignancy [3]. MPE can cause significant reduction in quality of life to patients due to symptoms of dyspnoea and cough [4].

Chemical pleurodesis is a recommended first line treatment in the management of MPE [3]. It is used to relieve symptoms and prevent the recurrence of MPE and has been in use for over 80 years, with an estimated 100,000 patients in the US undergoing pleurodesis every year [5]. A bedside chemical pleurodesis can cost up to $11,224 and an estimated inpatient annual expenditure of more than $5 billion in the US alone [6, 7]. Pleurodesis involves the administration of a chemical sclerosant within the pleural cavity, to cause pleural irritation leading to adhesion of the visceral and parietal pleura, hence, obliterating the pleural cavity and preventing the fluid accumulation [8, 9]. Talc is the preferred sclerosing agent of choice, which shows greatest efficacy and has a preferable safety profile compared to other agents, the most common side effects are mild and pleuritic chest pain and, a potential serious complication is the adult respiratory distress syndrome [10, 11]. Patients who undergo successful talc pleurodesis have a mean survival time of 11 months, opposed to 6.4 months in patients with failed pleurodesis [12]. Talc pleurodesis is carried out via two methods: talc poudrage (insufflation) performed during a thoracoscopy, and talc slurry, which is administered via a chest tube or a catheter. Efficacy of both methods are similar. However, some studies show talc poudrage as being more efficacious than talc slurry, due to greater success rates reported [3, 11, 13, 14]. This could be due to the direct visualisation of the pleural cavity and equal distribution of the talc within it. The decision for the use of poudrage or talc slurry depends on multiple factors, with talc slurry being less invasive than talc poudrage, in which the thoracoscopy procedure can cause complications, such as atelectasis, pneumonia, or respiratory failure [7, 14].

The success rate of talc pleurodesis for partial and complete response has been reported in most studies as 63-93% [3, 15, 16]. However, success rates as low as 43% and 50% have been reported as well [17, 18]. Multiple factors may affect the success rate of the talc pleurodesis. A low patient Karnofsky score status (a metric used to assess and quantify the functional performance of patients), age, gender, presence of pleural adhesions, pleural fluid pH, prolonged period between diagnosis of MPE and treatment, and use of systemic corticosteroids are associated with lower rates of successful pleurodesis [16, 19]. The type of the tumour associated with malignancy also influences success rates, with mesothelioma and lung cancer showing the lowest success rates [20]. The operator dependant nature of this procedure gives rise to the question whether injectability of the talc slurry may affect distribution. *Mager et al* observed that distribution of the talc slurry in patients was generally quite poor. 75% of the patients in the study had a dispersion of the talc covering less than 50% of the pleural cavity [21]. Furthermore, the talc slurry distribution was mostly dictated by the position of the tube, which is generally directed towards the basal part of the pleural cavity, for better fluid drainage [21].

This study aims to assess the distribution of the talc slurry within the pleural space via a chest tube, and to determine the potential for the use of a multidirectional two-tube method to improve the talc slurry distribution. Administration of viscous compounds can cause difficulty due to problems with injectability, which can affect the efficacy due to the force required to push them. [22]. This study also assesses the force required to push the talc slurry through a 14 Fr chest tube.

## Materials and Methods

### Talc Slurry Preparation

50ml of a sterile 0.9% normal saline (Baxter Healthcare Ltd, Norfolk, United Kingdom) was injected via a 50ml syringe (Terumo Europe N.V., Leuven, Belgium) into a vial containing 4g of sterile talc powder (STERITALC® F4, Novatech SA, La Ciotat, France). The contents of the vial were shaken to ensure all of the talc was suspended in the normal saline, and was then withdrawn into the syringe. For aiding visualisation in mechanical and distribution testing, 2 ml of green dye (water, tartrazine E102, green S E142, acetic acid, Goodalls Spices Green Colouring) was added to the talc slurry.

### Porcine thoracic bio-model

The thoracic bio-model consisted of a pig’s ribcage draped in 3M™ Ioban™ film, placed over a plastic scaffold. 3M™ Ioban™ film was placed over the plastic scaffold to mimic the visceral pleura, thus creating a simulated pleural cavity. A 14 Fr (Vygon, Ecouen, France) chest tube was inserted through the intercostal space and positioned within the simulated pleural cavity (Figure 1).

**Figure 1:**
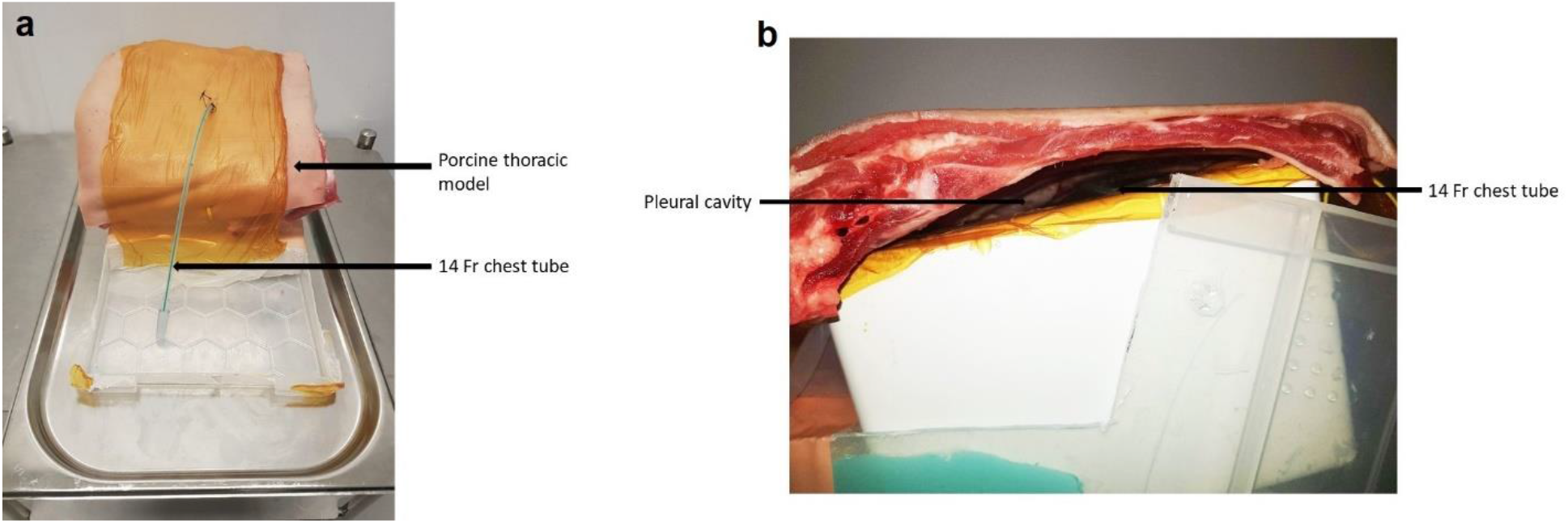
(a) Front view of porcine thoracic bio-model, draped in Ioban film (3M) with a 14 Fr chest tube inserted through the intercostal space. (b) Side view of the porcine thoracic bio-model showing the chest tube within the model pleural cavity.

### Mechanical Testing

Mechanical testing to determine the force required to push the talc slurry through a 14 Fr chest tube was performed using a Zwick mechanical testing machine (Z005 Zwick/Roelle, Germany) with a 50 Newton (N) load cell at a rate of 2.8 mm/s on a thoracic bio-model. A 14 Fr chest tube was connected to a 50ml syringe containing prepared talc slurry (Figure 2). The force required to push the talc slurry through the chest tube was recorded. Three replicates (n=3) were performed. The data is represented as mean +/- standard deviation (SD). Graphs were plotted using GraphPad Prism software.

**Figure 2:**
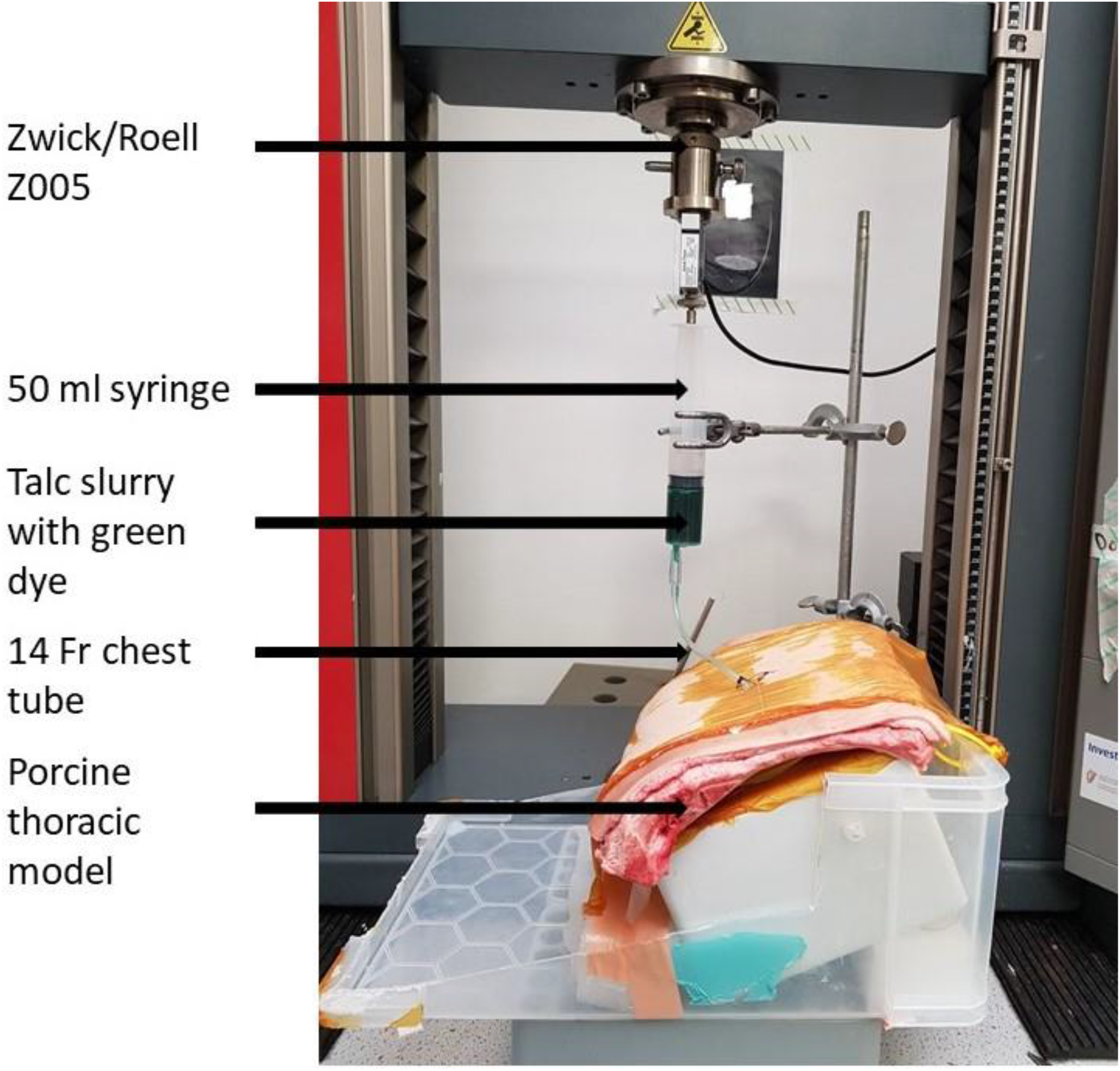
Setup of Zwick/Roelle z005 mechanical tester with 50ml syringe containing talc slurry with green dye connected via a 14 Fr chest tube to the porcine thoracic bio-model.

### Talc slurry distribution testing – human cadaver study

The distribution of the talc slurry within the pleural cavity was tested using human cadaveric models in supine position.

In cadaver 1, a 14 Fr chest tube with a length of 28cm was inserted using a trocar through the right 5^th^ intercostal space, anterior to mid-axillary line (Figure 4). 50ml of the prepared talc slurry was administered via the chest tube with 2 ml of green dye to aid visual assessment of its distribution. Following administration, a dissection was performed to remove the thoracic wall, using a surgical saw and rib shears. This allowed visualising the distribution of the talc slurry within the pleural cavity.

In cadaver 2, a second multidirectional two-tube method was used to test the distribution, and to compare with the first method. The aim was to investigate if this method leads to a more even distribution of the talc slurry than the method used in cadaver 1. The thoracic wall had already been removed from a previous dissection prior to the talc slurry administration. 25ml of the talc slurry was administered with green dye via a 14 Fr chest tube directed at two different sites (apically and basally) onto the visceral pleura of the right lung.

## Results

### Mechanical Testing

The force required to push the talc slurry through the 14 Fr chest tube in the porcine thoracic bio-model was found to be 11.36 +/- 2.79 N and remains relatively constant over time as shown in Figure 3 (a) and (b).

**Figure 3:**
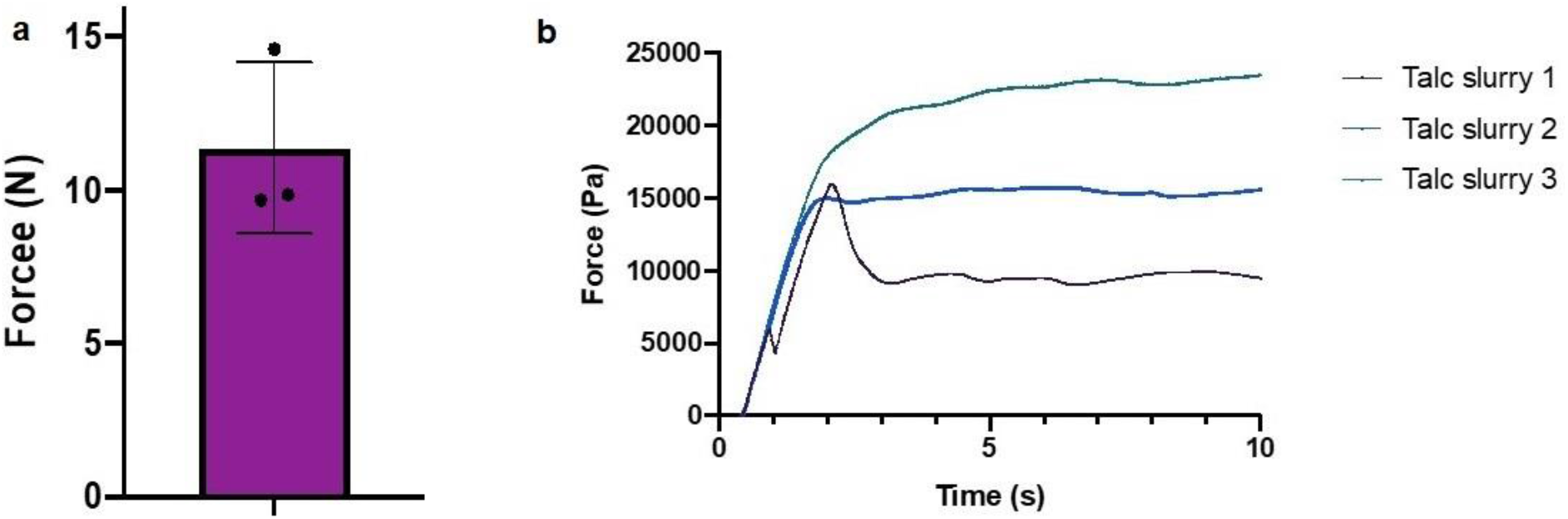
(a) Max force required to push the talc slurry through a 14 Fr chest tube was 11.36 N +/- 2.79 N. (b) Force required to administer talc slurry vs time.

**Figure 4:**
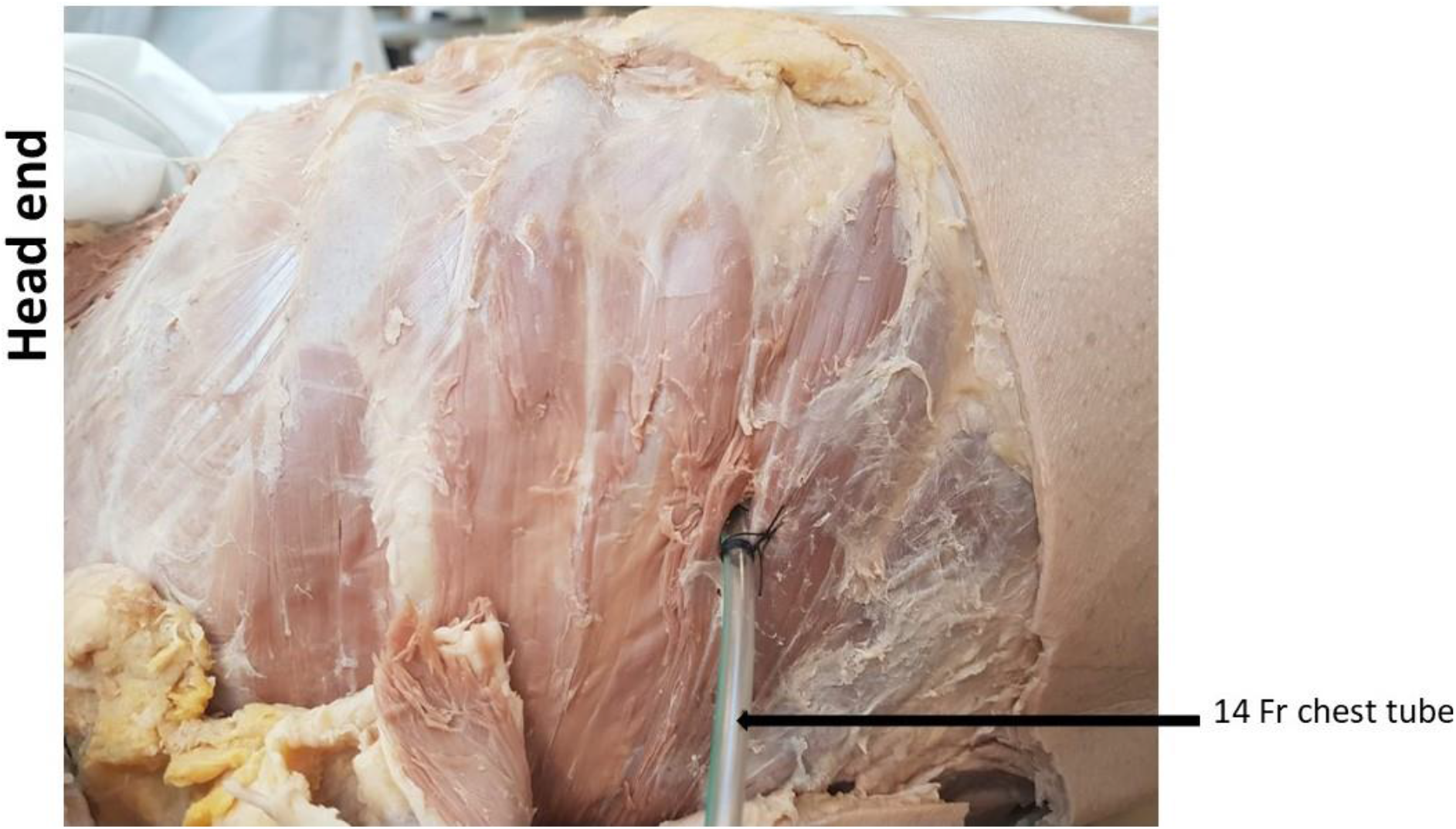
Cadaver 1: 14 Fr chest tube inserted at right 5th intercostal space of cadaver 1.

### Talc slurry distribution testing – cadaver study

The distribution of the talc slurry was assessed by visual inspection. In cadaver model 1, talc slurry was administered via a 14 Fr chest tube inserted within the pleural cavity through the right 5^th^ intercostal space, anterior to mid-axillary line, as is usually done in clinical practice (Figure 4).

Distribution of the talc slurry was unequal across the visceral pleura, with the majority of the talc slurry being concentrated at the diaphragmatic surface of the right lung, and little along the anterior surface as shown in Figure 5.

**Figure 5:**
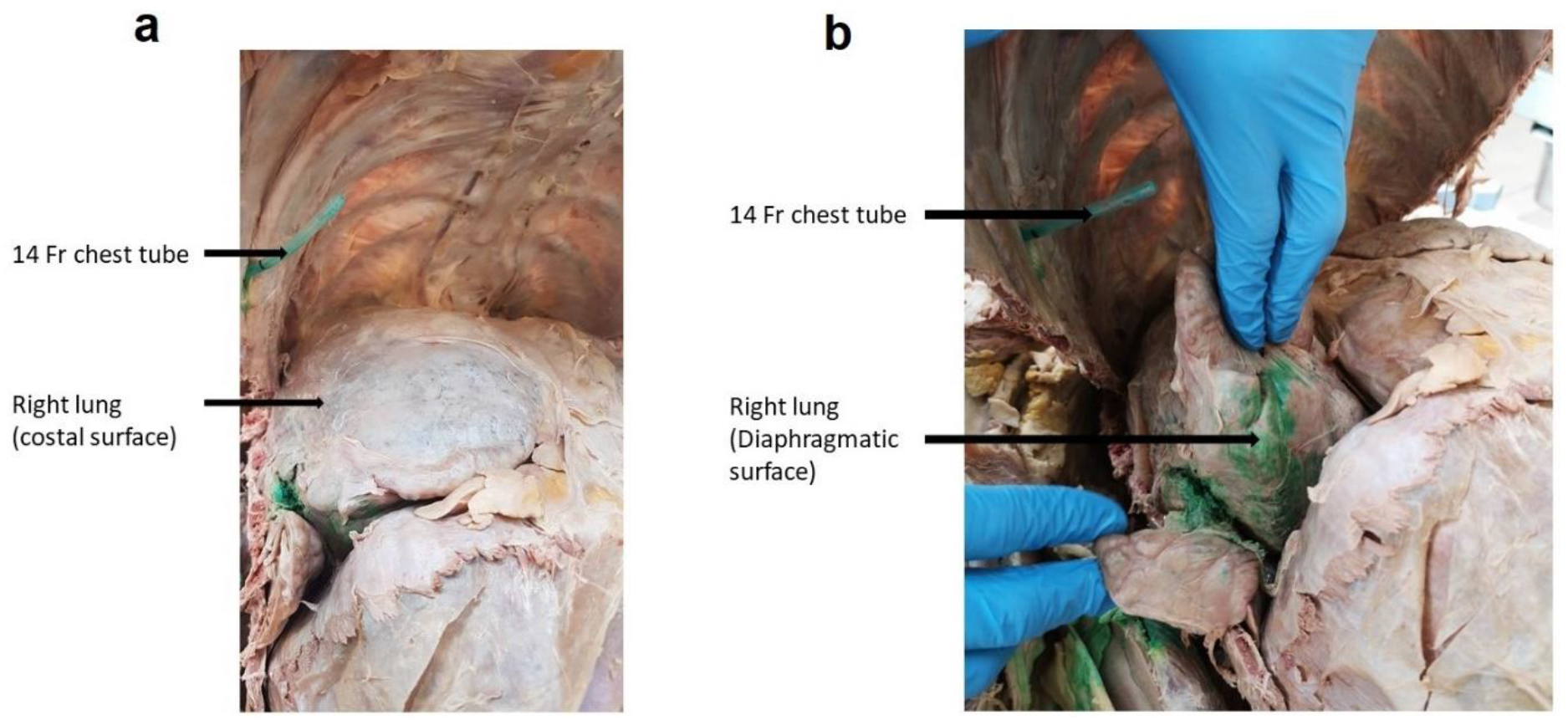
Distribution of the talc slurry (with green dye) within the pleural cavity of cadaver 1: (a) There is little talc deposited on anterior surface of the right lung. (b). Talc is mainly deposited along the diaphragmatic surface of the right lung.

In cadaver model 2, talc slurry was administered using the chest tubes in different directions, apically and basally, over the right lung. Talc covered more area across the visceral pleura, compared to the single tube method used in cadaver model 1. The talc slurry spread was noticeable more across the anterior surface of the lung, as pictured in Figure 6.

**Figure 6:**
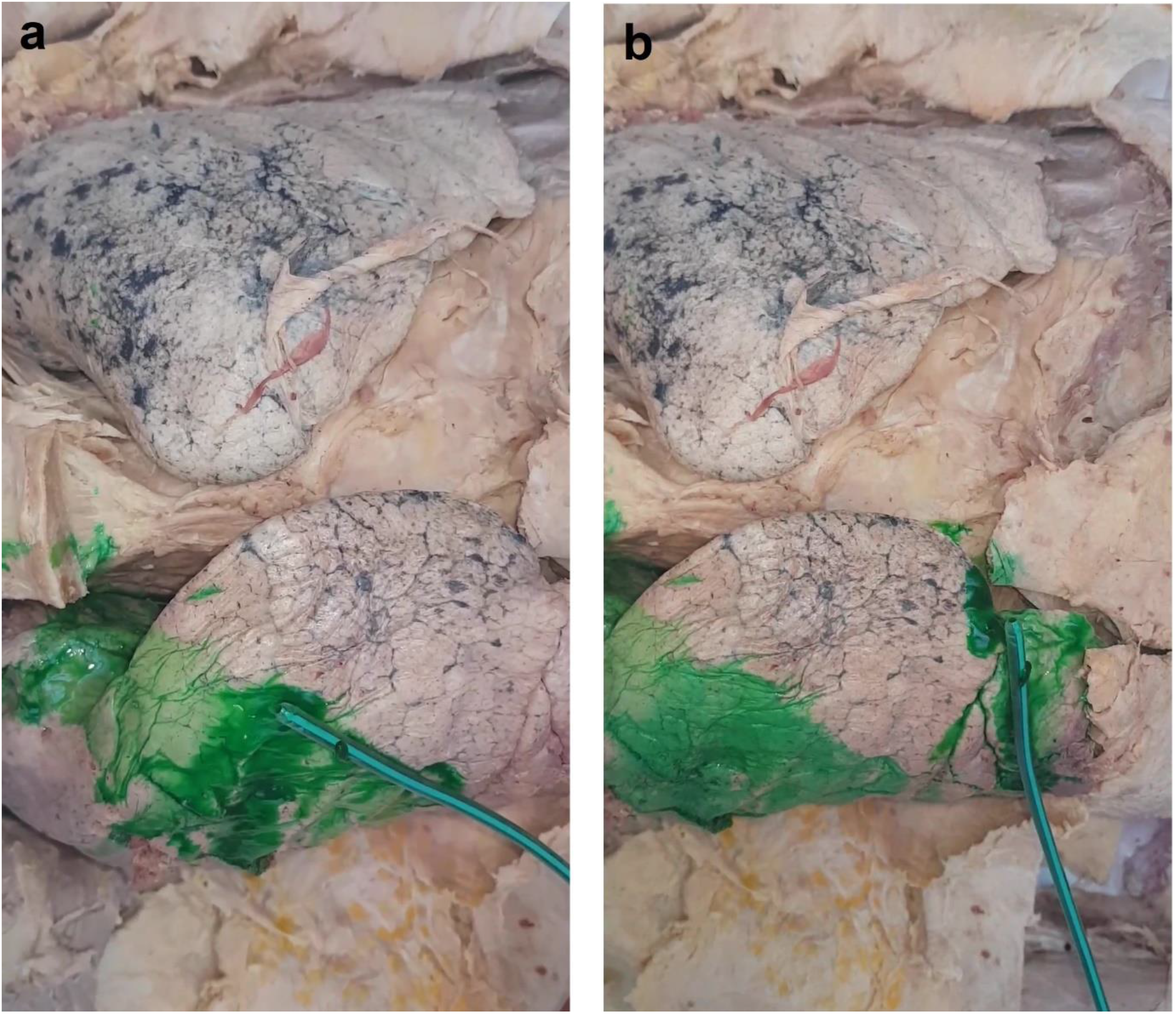
Better distribution of talc slurry over the right lung using the two-tube method on cadaver 2.

## Discussion

Mechanical testing using the porcine thoracic bio-model is effective in measuring the force required to push the talc slurry through a 14 Fr chest tube. Force required to inject a solution could affect the injectability of the solution, especially a viscous solution through a catheter [22]. A quick viscosity test suggests that the viscosity of the talc slurry is ∼ 4 centipoise (cP). For context, this viscosity is just slightly higher than that of full-fat milk at 4° C [23].The maximum force required to administer the talc slurry is ∼ 11.36 N This is well below the average pinch grip strength of males (70 N) and females (50 N) [24]. Furthermore, the force over time (Figure 3) shows that the force required to inject the talc slurry remains relatively constant over time. The low force required for administration means one would expect operators administering the talc slurry having no difficulty in mechanically pushing the talc slurry, those being able to deliver it at a steady rate.

This study shows that the talc slurry distribution within the pleural cavity is poor when administered via a basally placed chest tube. The majority of the talc slurry is concentrated around the diaphragmatic surface of the lung. In comparison, administration of the talc slurry using a multidirectional two-tube method (apical and basal) results in more even distribution across the anterior aspect of the lung. Mager *et al* [21] used radiolabelled talc and scintigraphy and found that the talc slurry had less than 50% coverage in the pleural cavity for 75% of total participants, and 15% of participants showing less than 25% talc slurry coverage [21].

Success rates reported in the literature for pleurodesis vary. This may be due to the result of an inconsistent definition of success and failure of pleurodesis being used across various studies. A meta-analysis outlined that failure rates are often defined only as a lack of radiological recurrence of MPE [25], while some are defined using patient based factors, such as recurrence of symptoms [26]. The timeframe for definition of success also differs. One definition is the absence of the MPE within 3 weeks after procedure [27], another is “no recurrence of MPE” (detected by thoracic ultrasound) and “no further procedures performed” for a minimum of 3 months [18]. Some define pleurodesis failure as whether a patient requires one of the following procedures within 90 days after the pleurodesis: removal of ≥100 mL of fluid during thoracocentesis; insertion of a chest tube for fluid management; insertion of an indwelling pleural catheter; or thoracoscopy of any kind [15]. This discrepancy between the success and failure definitions is a potential reason for such variance of success rates in the literature. In addition to this, some studies include a classification of “partial response”, the definition of which also varies but it can be understood as fluid re-accumulation greater than post-pleurodesis level but below the pre-pleurodesis level [28, 29]. However, there is deviation on whether the absence of symptoms of MPE is a requirement for a partial response. Aydogmus *et al* defined partial response as the absence of symptoms, while *Bielsa et al* described it as diminution of symptoms, rather than absence [28, 30]. Both papers include partial responses in their percentage for overall success rates. In contrast to this, Bhatnagar *et al* did not define or include partial response [15, 17]. Furthermore, there is a significant variance between study centres for talc slurry administration technique [25]. The variances include different chest tube sizes, length of chest tube clamping and timing of the removal of the chest tube [25].

The varying definitions may contribute to the statistical heterogeneity in the reported data. In view of these variances, the success rates of the talc pleurodesis should be looked at carefully. However, it is generally accepted that a successful pleurodesis is defined as achieving satisfactory apposition of the visceral and parietal pleural surfaces, often indicated by a fully expanded lung at the end of the procedure and no radiological or clinical recurrence of the effusion at long-term follow up [3, 10]. The contact area of the talc with the pleural surfaces should be extensive to induce inflammation to achieve successful and complete pleural adhesion [8, 21]. The inconsistency and large variance of success rates (of up to 50% difference) combined with the cost of pleurodesis means there is scope for further research in improving the success rates of the talc pleurodesis. This study has shown that a multidirectional two tube-method for administering the talc slurry can improve distribution of the talc within the pleural cavity, which could help in further increasing the success rates. A limitation to this study is that the visceral & parietal pleura are frequently stuck to each other in embalmed specimens. In order to mitigate this, the multidirectional two-tube method was tested in an already dissected cadaver. Another limitation is the inability to alter the position of the cadaver, post talc slurry administration. Although described in some studies, rotation of the patient is not found to be associated with better distribution of the sclerosing agent [21]. Further research on a multidirectional two-tube method would be worth investigating to see if it is efficacious in improving the success rates of the chemical pleurodesis.

## Conclusion

The force required to administer the talc slurry through a syringe with a 14 Fr chest tube is relatively low to human pinch grip strength. The distribution of the talc slurry within the pleural cavity using a single tube method, as per current practice, is poor. A multidirectional two-tube method may provide a comparably better distribution of the talc slurry within the pleural cavity. This method has a scope to improve the success rates for talc pleurodesis.

## Data Availability

All data is already included in the paper.

## Acknowledgement

Mr. Miroslav Voborsky from Surgical simulation lab in RCSI University of Medicine and Health Sciences for his help in creating the porcine thoracic bio-model.

## Declarations

### Ethics approval

Approved by the Museum committee within Department of Anatomy and Regenerative Medicine, RCSI University of Medicine and Health Sciences.

### Consent to participate and consent for publication

Not applicable.

### Author contributions

**Eoin Campion:** Acquisition and analysis of the data, drafting the manuscript. **Saad I. Mallah**: Drafting the manuscript, resources. **Maimoona Azhar:** Methodology, revising the draft critically, editing and final approval of the manuscript. **Dara O’Keeffe:** Methodology, critical reviewing of the manuscript. **Aamir Hameed**: Conceptualisation, critical reviewing and final approval of the manuscript.

## Abbreviations

MPE: Malignant pleural effusion
N: Newton
Fr: French
cP: Centipoise

